# Warning labels for non-nutritive sweeteners: impacts on Chilean parents’ choices and perceptions

**DOI:** 10.64898/2025.12.22.25342864

**Authors:** Aline D’Angelo Campos, Lindsey Smith Taillie, Allison Sylvetsky, Natalia Rebolledo

## Abstract

**Background:** In 2016, Chile mandated that products high in sugar carry warning labels. Following this policy, use and consumption of non-nutritive sweeteners (NNS) increased, raising concerns about intake among children. Chile is now considering adding a new warning to its labeling system stating: “CONTAINS NON-NUTRITIVE SWEETENERS, AVOID CONSUMPTION AMONG CHILDREN.” However, this and other NNS labeling approaches – e.g., Colombia’s “CONTAINS NON-NUTRITIVE SWEETENERS” stop signs – have never been empirically tested.

**Methods:** In 2025, Chilean parents of children ages 2-14 (n=3,523) completed an online experiment. Participants were randomly assigned to see either no NNS label (control), Chile’s proposed NNS label, or Colombia’s existing NNS label on NNS-containing products. Participants saw four products (one without added sweeteners and three sweetened with either sugar, NNS, or both) and were asked to select one for their child, to identify NNS-containing products, and, afterwards, to report which labels they saw. Next, participants saw NNS-containing products alone and rated the NNS labels’ perceived message effectiveness (PME) and products’ healthfulness. Lastly, participants rated their intentions to limit their children’s future NNS intake. We report average differential effects (ADEs) compared to control in percentage points (pp) or on scales ranging from 1 (low) to 5 (high).

**Results:** Both the Chilean (ADE=67pp, p<0.001) and the Colombian (ADE=69pp, p<0.001) labels led to higher identification of NNS-containing products. Both labels led to lower selection of NNS-containing products (Chilean label ADE range=-22pp,-35pp; Colombian label ADE range=-22,-33pp; all p<0.001) and to higher selection of products without added sweeteners (Chilean label ADE range=20,35pp; Colombian label ADE range=22,31pp; all p<0.001). Neither label led to higher selection of sugar-sweetened products. Afterwards, 63% of participants in the Chilean NNS label condition and 84% in the Colombian NNS label condition reported seeing NNS labels (p<0.001). Lastly, both labels led to higher PME (Chilean label ADE=1.53; Colombian label ADE=0.92, both p<0.001), lower product healthfulness perceptions (Chilean label ADE=1.43, Colombian label ADE=0.81, both p<0.001), and higher intentions to limit children’s NNS intake (Chilean label ADE=0.66, Colombian label ADE=0.17, both p<0.001).

**Conclusions:** NNS labels greatly improved parents’ NNS identification and led to healthier product choices for their children, showing promise for curbing NNS intake in this vulnerable population.

## Introduction

Non-nutritive sweeteners (NNS), which are food additives commonly used as sugar substitutes, have been at the center of controversy for some time due to potential health concerns. While studies show mixed results for the association between NNS intake and health outcomes in animal and human models, with some indicating potential gut microbiome alterations and glucose intolerance.^1–4^ Although the translation of these findings to human health under usual consumption levels remains uncertain, upon reviewing the available evidence in 2023, the World Health Organization conditionally recommended against using NNS for weight control and noncommunicable disease prevention.^5^ Additionally, NNS consumption can be particularly concerning for children. Sweet taste preferences develop in early childhood, raising concerns that exposure to NNS at young ages might reinforce preferences that could lead to a lifelong overconsumption of sweet foods and beverages.^6–8^ Children can also be at higher risk of exceeding safe NNS intake limits due to their lower body weight.

In 2016, the Chilean government implemented a pioneer policy mandating front-of-package warning labels (FOPWLs) on products high in energy, sugars, saturated fats, and sodium. Additionally, products that qualified to carry FOPWLs were banned from schools and became subject to restrictions in child-directed marketing.^9^ After the initial implementation of this policy, a large-scale reformulation of packaged products took place, likely in an effort by manufacturers to bring nutrient contents below the FOPWL thresholds. Overall, the percentage of products required to display FOPWLs fell from 51% to 44% after implementation of the policy, with especially meaningful decreases in the percentage of products qualified as high in added sugars.^10^ However, simultaneously, the use of NNS in packaged products increased from 37.9% to 43.6%.^11^ As a result, children’s NNS intake also rose: NNS consumption prevalence rose from 77.9 to 92% among preschool children, with significant increases in the intake of four different types of NNS.^12^ Given this possible unintended consequence of FOWPL and related policies, the Pan American Health Organization has included NNS in its nutrient profile model as a food additive of concern,^13^ and countries to later adopt FOPWL policies similar to Chile’s – including Mexico, Argentina, and Colombia – have implemented different types of NNS warning labels as part of their FOPWL systems. However, none of these NNS warning labels have been empirically tested to date.

Amid rising concerns, in June of 2024, the Chilean Ministry of Health proposed modifying its labeling system to include a rectangular label stating: “CONTAINS NON-NUTRITIVE SWEETENERS, AVOID CONSUMPTION AMONG CHILDREN.”^14^ This label would emulate the language and design of Mexico’s and Argentina’s NNS warning labels, but would differ considerably from Chile’s existing FOPWLs, which are octagonal (resembling stop signs) and use more concise language (e.g., “HIGH IN SUGAR”). Thus, questions remain about whether emulating Colombia’s NNS warning label, which is octagonal and simply states “CONTAINS NON-NUTRITIVE SWEETENERS” might be a more effective strategy. Additionally, there are concerns that the introduction of NNS warning labels could backfire and lead parents to choose products with added sugars in attempts to avoid NNS in their children’s diets.

This study aims to evaluate and compare the impact of two different types of NNS warning labels (Chile’s proposed label and Colombia’s existing label) on Chilean parents’ ability to identify products containing NNS, their product choices for their children, and their product perceptions and future behavioral intentions.

## Methods

### Participants

In July and August 2025, we recruited an online convenience sample of Chilean parents through the panel company Netquest. Participants were eligible if they were at least 18 years old, resided in Chile, and had a child between 2 and 14 years old who had never been diagnosed with diabetes, prediabetes, or insulin resistance. We used quotas to ensure that approximately 50% of the sample had an educational attainment of high school or less. The University of North Carolina at Chapel Hill Institutional Review Board approved this study (#24-2896). All participants provided electronic informed consent. The study design, measures, hypotheses, and analytic plan were registered before data collection on ClinicalTrials.gov (NCT06842693).

### Procedures

Participants completed an online survey programmed in Qualtrics software and translated to Spanish by native Chilean Spanish speakers. Using the Qualtrics randomizer function, participants were randomized into one of three NNS labeling conditions using a 1:1:1 allocation ratio: control, Chile NNS label, or Colombia NNS label. In the control condition, no label signaled the presence of NNS. In the Chile NNS label condition, NNS-containing products carried the label currently under consideration in Chile (rectangular “CONTAINS NON-NUTRITIVE SWEETENERS, AVOID CONSUMPTION AMONG CHILDREN”). In the Colombia NNS label arm, NNS-containing products carried the label currently used in Colombia (octagonal “CONTAINS NON-NUTRITIVE SWEETENERS”). Notably, the original Spanish term used in both types of NNS labels (and in our outcome measures) was “*edulcorantes” –* a technical term that encompasses both artificial and natural types of NNS. This term does not have a direct English translation, so we use simply “non-nutritive sweeteners” for the purposes of this study.

The experiment included tasks using images of fictitious food products to avoid the influence of existing brand preferences. Products included labels according to participants’ randomly assigned labeling condition. Due to limited screen space, the labels were small and somewhat difficult to read on the product packages, so we also displayed enlarged labels in call-out boxes next to each product – an approach used in previous studies^15–18^ as well as by real Chilean online grocery stores.

In the first task, participants saw three sets of products in random order and completed selection tasks for each set (described in detail under *Measures*). Each set corresponded to a product category (i.e., fruit drinks, yogurts, and breakfast cereals) and contained four products shown in random order: one product without added sweeteners, one sugar-sweetened product, one NNS-sweetened product, and one sugar-plus-NNS-sweetened product. Regardless of NNS labeling condition, sugar-sweetened products in this task carried an octagonal “HIGH IN SUGAR” label, reflecting Chile’s current labeling policy. Within each set, all products had the same flavor and similar package design. On the package, the NNS-sweetened product stated “no added sugar” and the NNS-plus-sugar-sweetened product stated “with 50% less calories,” reflecting common claims on these sorts of products (Figure 1).

**Figure 1.**
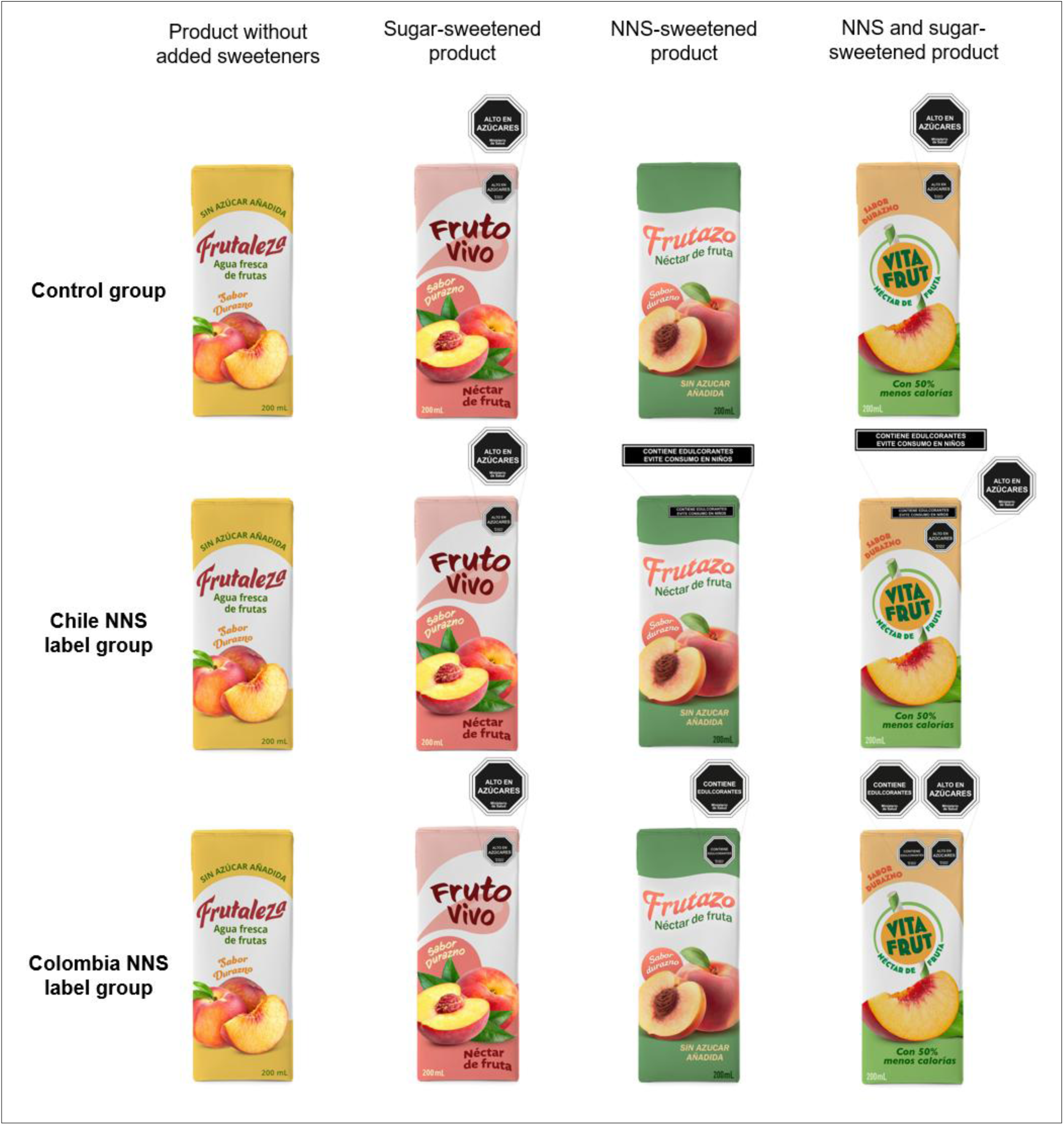
Example set of products used in task 1 by label type

The second task consisted of single product assessments. Participants saw a chocolate-flavored milk and a cereal bar, displayed one at a time in random order, and completed measures of product and label perceptions for each product (also described in detail under *Measures*). Both products were NNS-sweetened and stated “no added sugar.” In the control condition, products in this task carried a neutral barcode label so that participants could respond to measures that referenced a label, following an approach established in prior studies.^15,19–21^ In the Chile NNS label and Colombia NNS label conditions, products carried the same NNS labels as in the first task (Figure 2).

**Figure 2.**
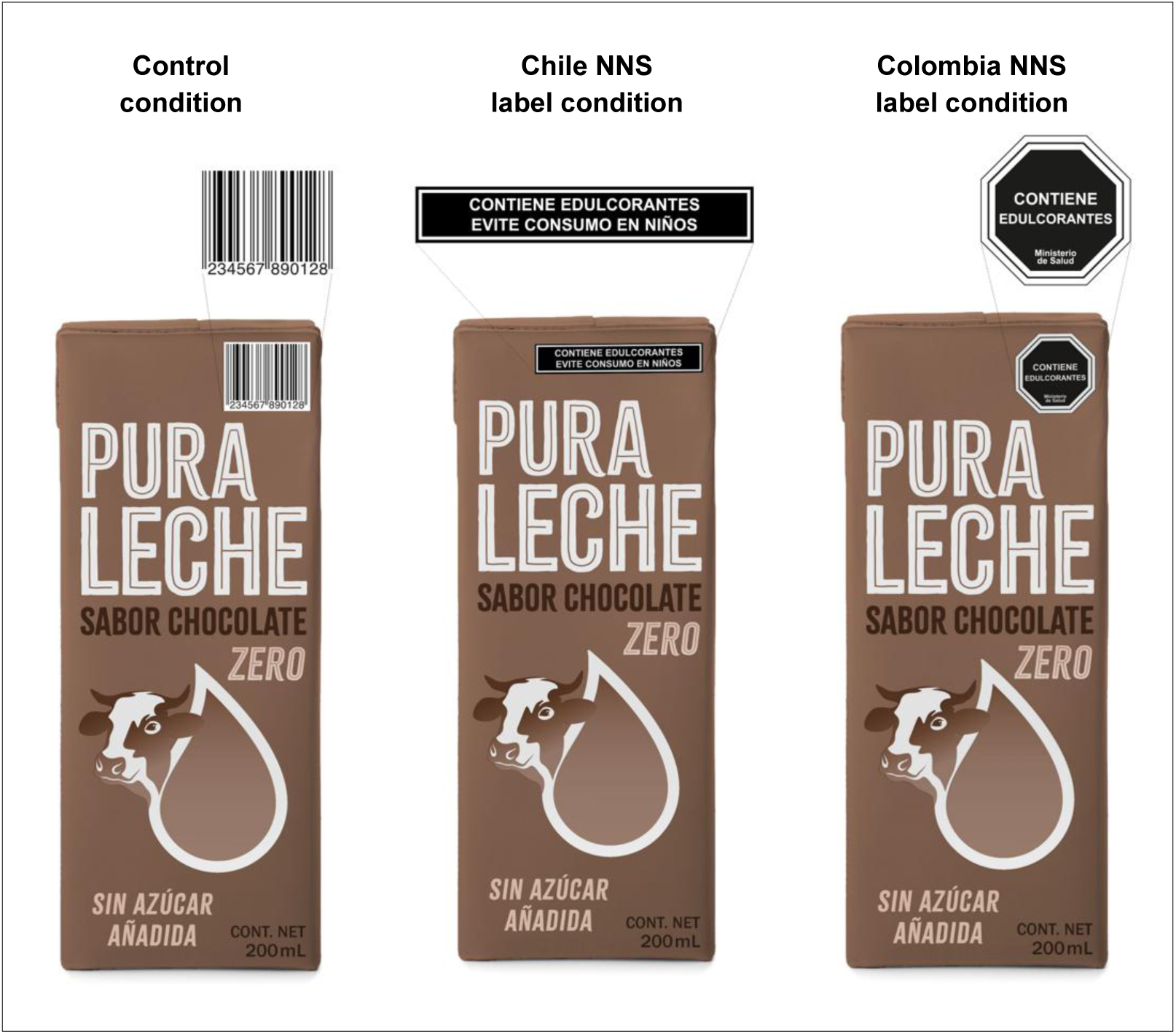
Example product used in task 2

### Measures

In the first task, participants were asked to select one product that they would purchase for their child from each set of products. Next, participants were asked to identify all products that contained NNS from each set of products. After completing these measures for all sets of products, participants were asked whether they recalled seeing front-of-package labels on the products during the task. Those who replied “yes” were further asked what the labels that they recalled seeing were about. Response options were non-exclusive and included sugar, NNS, calories, sodium, saturated fats, caffeine, and trans fats. Calories, sodium, and saturated fat labels were not displayed during the experiment but are part of Chile’s existing FOPWL system, while caffeine and trans fats labels were neither displayed during the experiment nor part of Chile’s existing FOPWL system.

In the second task, we measured the NNS labels’ perceived message effectiveness (PME). This measure comprised three items, assessing: (1) how much the label discouraged participants from wanting to buy products with NNS for their child, (2) how much the label made participants concerned about the health effects for their child of consuming products with NNS, and (3) how much the label made buying products with NNS for their child unpleasant. PME has been shown to predict behavior change in the context of warning label exposure.^22,23^ Responses were assessed on a 5-point scale from “not at all” (coded as 1) to “very much” (coded as 5). Next, we measured products’ perceived healthfulness (i.e., how good or bad for their child’s health it would be to consume the product every day). The 5-point response scale ranged from “very bad” (1) to “very good” (5). We also measured products’ relative healthfulness (i.e., how healthful participants believed the product to be compared to a version of the same product sweetened with sugar). The 5-point response scale ranged from “much less healthy” (1) to “much healthier” (5).

Lastly, after both tasks, we assessed how much participants intended to limit their child’s non-nutritive sweetener consumption over the next week on a 5-point scale ranging from “not at all” (1) to “completely” (5). Participants also answered questions about their prior experiences with NNS and standard sociodemographic characteristics.

### Analyses

We excluded participants who completed the survey implausibly quickly (i.e., in less than one-third of the median completion time), who completed less than 90% of the survey, or who had reCAPTCHA scores below 0.5.

To analyze results from the first experimental task, we coded outcomes as dichotomous. When asked to identify NNS-containing products, we coded whether participants selected both NNS-containing products and no others (i.e., correct identification) or not (i.e., incorrect identification), as our goal was to assess whether participants could reliably identify NNS presence across multiple product formulations. When asked to select a product to purchase for their child, we coded whether participants (a) selected one of the NNS-sweetened products or not, (b) selected the product without added sweeteners or not, and (c) selected one of the sugar-sweetened products or not. We then used logistic mixed-effects regression models to analyze the effects of the labels on each of these outcomes, treating the intercept as random to account for repeated measures within participants. Models regressed each outcome on indicator variables for the labeling conditions, product categories (i.e., fruit drink, yogurt, or breakfast cereal), and the interactions between labeling conditions and product categories. Post model estimation, we conducted Wald tests to assess the joint statistical significance of the interaction terms. The interactions were *not* jointly significant when modeling correct identification of NNS-containing products, so we dropped them from the final model for this outcome. Given that predicted probabilities of the outcome might still differ in non-linear models without significant interactions,^24^ we conducted additional post-estimation Wald tests to assess the joint significance of differences in predicted probabilities across product categories. These tests also did not yield significant results, so we report average differential effects (ADEs) on correct identification of NNS-containing products (i.e. differences in the predicted probability of the outcome between labeling conditions) across product categories. Alternatively, for the models examining selection of NNS-sweetened product, selection of product without added sweeteners, and selection of sugar-sweetened product, the interaction terms were jointly significant - so we kept them in the final models and report ADEs separately for each product category. Alternatively, to analyze the impacts of the labels on recall, which was measured only once after the selection tasks, we used logistic models, regressing each nutrient or ingredient provided as a response option on labeling conditions.

To analyze results from the second experimental task, we first verified that Cronbach’s alpha was sufficient (>0.7) and averaged participants’ scores across the three PME items for each product. Next, we used linear mixed-effects regression models to analyze the effects of the labels on PME, perceived healthfulness, and relative healthfulness, treating the intercept as random to account for repeated measures within participants. Models regressed each of these outcomes on indicator variables for the labeling conditions, product categories (i.e., chocolate milk or cereal bar), and their interactions. Post model estimation, we conducted Wald tests to assess the joint statistical significance of the interaction terms. None of the models presented jointly significant interactions, so we dropped the interaction terms in the final models and present ADEs (i.e. differences in the predicted mean outcome between labeling conditions) across product categories. Alternatively, to analyze the impacts of the labels on participants’ intentions to limit their child’s NNS consumption, which was measured only once after all product exposures, we used linear models, regressing the outcome on labeling conditions.

Lastly, we examined whether the effects of the labels on correct identification of NNS-containing products, selection of NNS-sweetened product, and selection of unsweetened product (outcomes from the first experimental task) differed by participants’ gender, educational attainment, and self-reported use of tabletop NNS. We used the latter as a proxy for participants’ intentional use of NNS, assuming that those who voluntarily add NNS to foods or beverages demonstrate more deliberate intentions to use NNS than those who only consume NNS incidentally through ready-to-eat or ready-to-drink products, which may contain NNS without their awareness. We dichotomized the moderator variables to maximize power and simplify interpretations. We used mixed-effects regression models, regressing outcomes on indicator variables for the labeling conditions, the moderator (one moderator per model), and their interactions. Due to significant interactions between labeling conditions and product categories in the main models for selection of NNS-sweetened product and selection of unsweetened product, we fit separate mixed-effects moderation models for each product category (i.e., fruit drink, yogurt, and breakfast cereal) for these outcomes.

Analyses included all participants according to the trial arm to which they were randomized. We used complete case analysis, resulting in the exclusion of 3 participants from some of our primary outcomes. Analyses were conducted using Stata/MP version 19.5 with a two-sided critical alpha of 0.05. We made some deviations from the registered analytic plan: (1) we did not register our plan to exclude participants with reCAPTCHA scores below 0.5, but decided to exclude such participants per advice from the Qualtrics platform; (2) we registered our plan to only report label noticing and label recall descriptively, but decided to conduct statistical testing on label recall post-hoc given its relevance to the study’s objectives; and (3) we registered that we would use mixed-effects models for all outcomes, but used logistic and linear regressions models with only fixed parameters to analyze effects on label recall and intentions to limit one’s child’s NNS intake because these outcomes did not include repeated measures, and (4) we did not our plan to examine moderation by NNS tabletop use, but decided to include this analysis post-hoc to better understand whether the effects of the NNS labels may differ among individuals who use NNS intentionally.

## Results

The final analytic sample included 3,523 participants (Figure 3). Participants’ mean age was 39.6 and 70% identified as women. Around 44% had an educational level of high school or less, 11% identified as Indigenous, and 59% considered themselves overweight or very overweight. Participants’ children’s mean age was 8.9. Around 19% of participants considered their child overweight or very overweight, and 27% reported having been previously told by a health care provider that their child needed to lose weight (Table 1).

**Figure 3.**
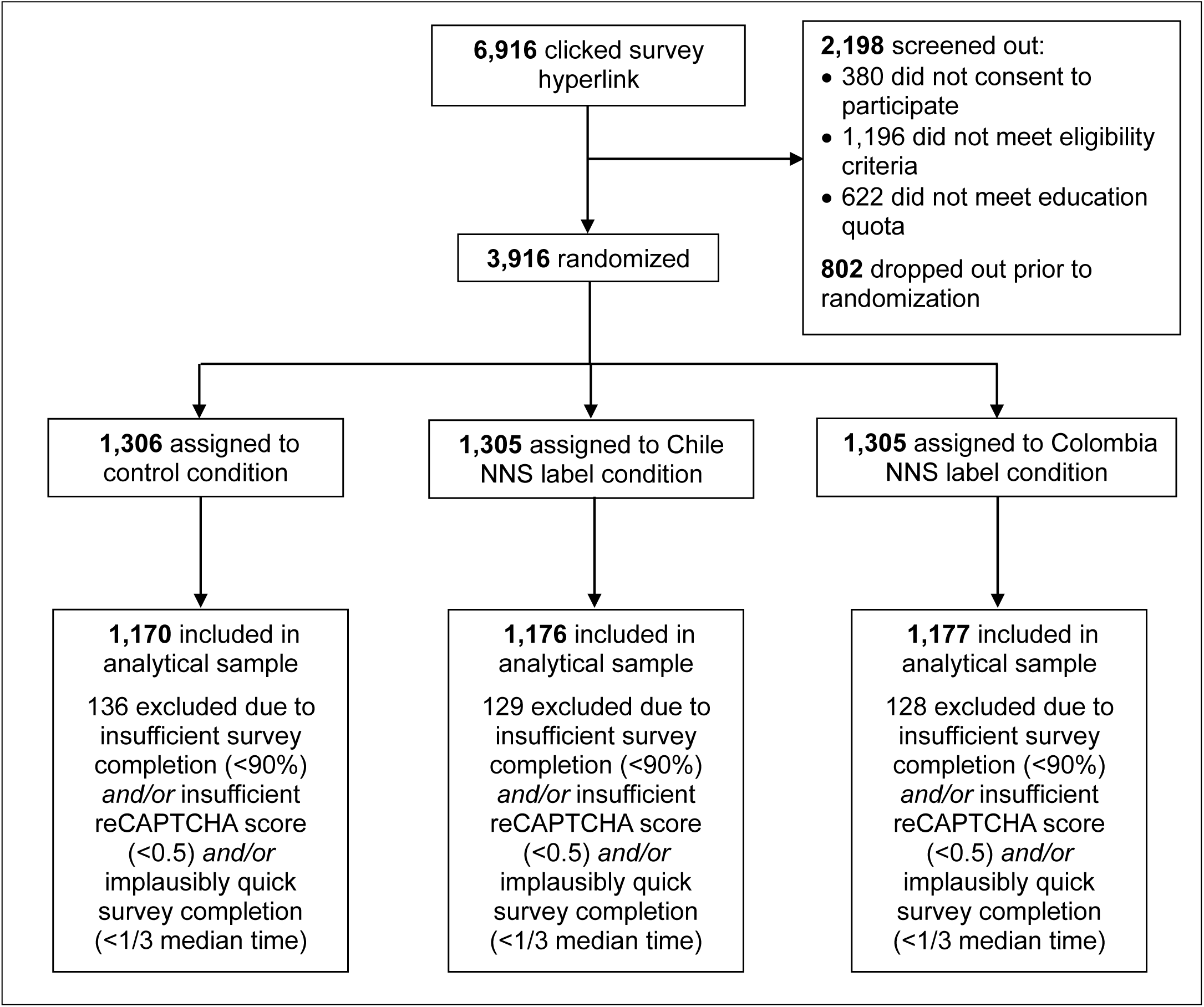
Participant flow diagram (CONSORT)

**Table 1.** Participant characteristics (n=3,523)

| **Characteristics** | **n** | **%** |
| --- | --- | --- |
| **Gender** |  |  |
| Man | 1,050 | 30% |
| Woman | 2,470 | 70% |
| Other | 3 | <1% |
| **Education** (n=3,509) |  |  |
| None, kindergarten, or basic education | 107 | 3% |
| High school | 1,435 | 41% |
| Higher-level technician (1-3 years) | 836 | 24% |
| Professional (4+ years) | 919 | 26% |
| Post-graduate | 212 | 6% |
| **Ethnicity** (n=3,506) |  |  |
| Indigenous | 374 | 11% |
| **Region** (n=3,496) |  |  |
| Metropolitana | 1,774 | 51% |
| Valparaíso | 391 | 11% |
| Biobío | 246 | 7% |
| North (Antofagasta, Atacama, Arica y Parinacota, Coquimbo, Tarapacá) | 271 | 8% |
| Central-West (O'Higgings, Ñuble, Maule) | 365 | 10% |
| South (Aysén, La Araucanía, Los Lagos, Los Ríos, Magallanes) | 449 | 13% |
| **Nationality** (n=3,513) |  |  |
| Chilean only | 3,158 | 90% |
| Chilean and another | 97 | 3% |
| Other | 258 | 7% |
| **Perceived weight** (n=3,500) |  |  |
| Very underweight | 14 | <1% |
| Underweight | 70 | 2% |
| About the right weight | 1,335 | 38% |
| Overweight | 1,833 | 52% |
| Very overweight | 248 | 7% |
| **Ever diagnosed with** |  |  |
| Diabetes (n=3,506) | 575 | 16% |
| Hypertension (n=3,506) | 545 | 16% |
| High cholesterol (n=3,506) | 675 | 19% |
| Cardiovascular disease (n=3,506) | 60 | 2% |
| **Perceived income adequacy**<sup>~</sup> (n=3,433) |  |  |
| Somewhat or very difficult | 2,454 | 71% |
| Neither difficult nor easy | 725 | 21% |
| Somewhat or very easy | 254 | 7% |
| **Frequency of FOPL use** (n=3,432) |  |  |
| Never or rarely | 690 | 20% |
| Sometimes | 1,075 | 31% |
| Often or all the time | 1,667 | 49% |
| **Sources on NNS intake in the last 7 days** (n=3,513) |  |  |
| Tabletop NNS | 1,821 | 52% |
| Sugar-free soda | 1,799 | 51% |
| Sugar-free fruit drinks | 1,525 | 43% |
| Sugar-free flavored yogurts | 1,449 | 41% |
| Sugar-free flavored water | 1,230 | 35% |
| Sugar-free flavored milk | 1,138 | 32% |
| Sugar-free sports drinks | 347 | 10% |
| Sugar-free energy drinks | 307 | 9% |
| None | 297 | 8% |
| **NNS recommendation from health care provider** (n=3,517) |  |  |
| Recommendation to consume NNS | 779 | 22% |
| Recommendation to avoid NNS | 876 | 25% |
| No recommendation | 1,862 | 53% |
| **NNS-related effort in the last year** |  |  |
| Made effort to consume more NNS | 314 | 9% |
| Made effort to consume less NNS | 2,049 | 58% |
| Did not make any NNS-related effort | 1,148 | 33% |
| **Best term to describe NNS** |  |  |
| Artificial sweeteners | 1,289 | 37% |
| Sweeteners | 936 | 27% |
| "Edulcorantes" | 447 | 13% |
| Sugar substitutes | 378 | 11% |
| Non-nutritive sweeteners | 169 | 5% |
| Non-caloric sweeteners | 154 | 4% |
| Diet sweeteners | 113 | 3% |
| Other | 33 | 1% |
| **Child's sex** |  |  |
| Masculine | 1,779 | 50% |
| Feminine | 1,744 | 50% |
| **Child's perceived weight** (n=3,503) |  |  |
| Very underweight | 16 | <1% |
| Underweight | 188 | 5% |
| About the right weight | 2,646 | 76% |
| Overweight | 625 | 18% |
| Very overweight | 28 | 1% |
| **Health care provider says child needs to lose weight** (n=3,503) | 931 | 27% |
|  | **Mean** | **SD** |
| **Age** | 39.6 | 7.8 |
| **Child age** | 8.9 | 3.6 |
| **Household size** (n=3,432) | 4.1 | 1.4 |

~Measured as perceived difficulty of making ends meet without going into debt
Note: Sample size is provided for specific variables when data was not available for the entire analytic sample for that variable

Regarding previous experiences with NNS, only 8% of participants did not report consuming any of the common sources of NNS assessed (including tabletop NNS and sugar-free products) in the last 7 days. Similar percentages of participants reported having previously received recommendations from health care providers to consume (22%) or avoid (25%) NNS. Around 58% reported making an effort to reduce their NNS intake in the last year. Participants’ most commonly preferred term to refer to NNS was “artificial sweeteners” (37%), followed by simply “sweeteners” (27%), and by the technical term “*edulcorantes*” (13%; Table 1).

### Selection tasks

Compared to participants in the control condition, those in the Chile NNS label condition were much more likely to correctly identify both NNS-containing products (67% vs. 0.5%, p<0.001), as were those in the Colombia NNS label condition (70% vs. 0.5%, p<0.001). Comparing between the two types of NNS labels, correct identification of NNS-containing products was slightly more likely among participants in the Colombia NNS label condition than among participants in the Chile NNS label condition (70% vs. 67%, p=0.002; Figure 4, Figure S1, Table S1).

**Figure 4.**
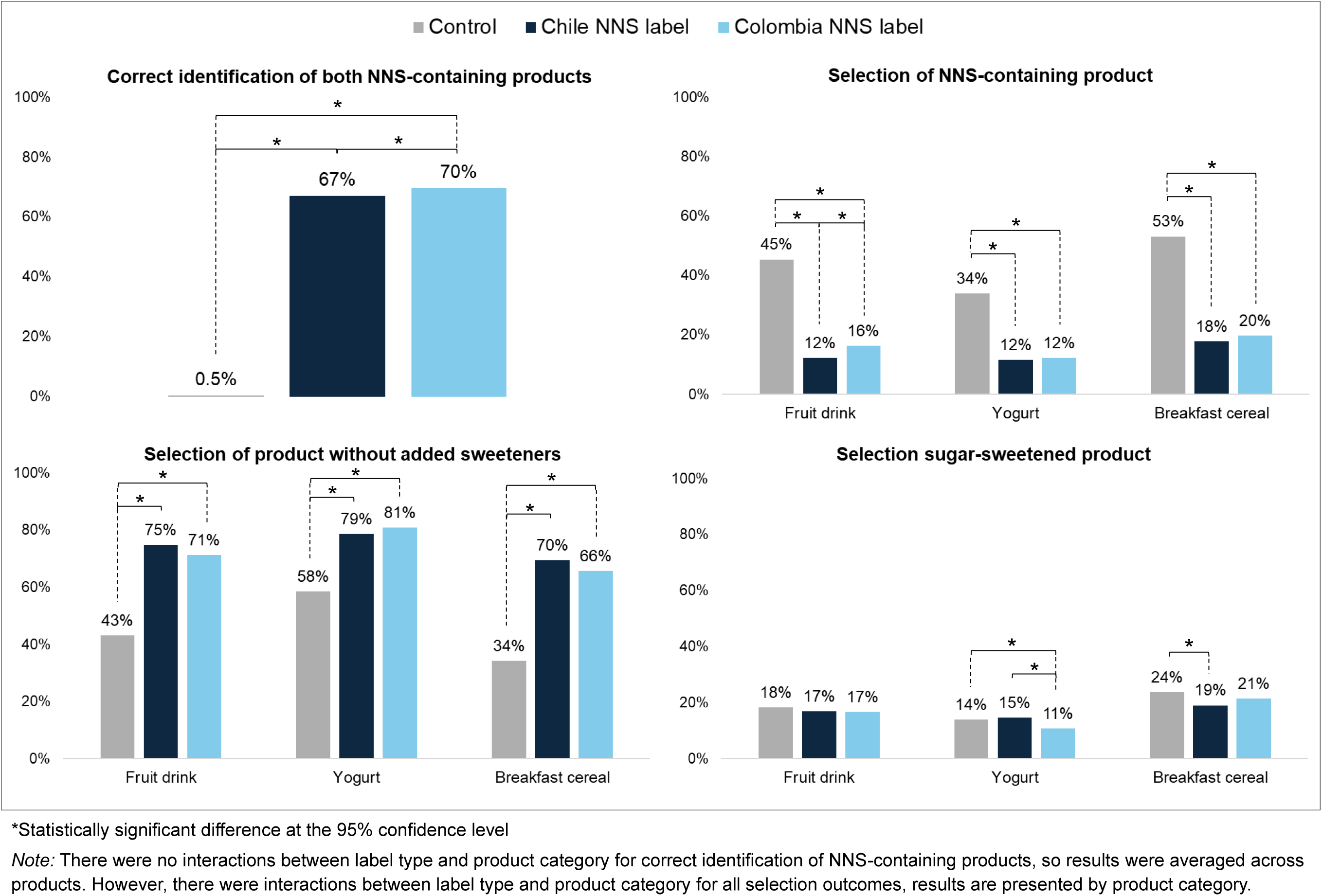
Predicted probability of correctly identifying both NNS-containing products (n=3,523) and of selecting one of the NNS-sweetened products, the unsweetened product, or one of the sugar-sweetened products to purchase for their child (n=3,520), by label type

Selecting a product containing NNS for hypothetical purchase was less likely among participants in the Chile NNS label condition (range: 12%-18%) and in the Colombia NNS label condition (range: 12%-20%) than among those in the control condition (range: 34%-53%) for the three product categories examined (p<0.001 for all comparisons to control). There were no significant differences in the likelihood of selecting a yogurt or a breakfast cereal containing NNS between the Chile NNS label condition and the Colombia NNS label condition (both p>0.05), but selecting a fruit drink containing NNS was slightly more likely in the Colombia NNS label condition than in the Chile NNS label condition (16% vs. 12%, p=0.01; Figure 4, Figure S2, Table S1).

Selecting the product without added sweeteners for hypothetical purchase was more likely among participants in the Chile NNS label condition (range: 70%-79%) and in the Colombia NNS label condition (range: 66%-81%) than among those in the control condition (range: 34%-58%) for all three product categories examined (p<0.001 for all comparisons to control). There were no significant differences in the likelihood of selecting the product without added sweeteners between the Chile NNS label condition and the Colombia NNS label condition for any of the three product categories examined (all p>0.05; Figure 4, Figure S2, Table S1).

There were no significant differences in the likelihood of selecting a sugar-sweetened fruit juice or hypothetical purchase between any of the arms (p>0.05 for all comparisons). The likelihood of selecting a sugar-sweetened yogurt did not differ between participants in the control condition and in the Chile NNS label condition (p>0.05), but both were more likely to select a sugar-sweetened yogurt than participants in the Colombia NNS label condition (respectively: 14% vs. 11%, p=0.02; 15% vs. 11%, p=0.003). Participants in the Chile NNS label condition were less likely to select a sugar-sweetened breakfast cereal than participants in the control condition (24% vs. 19%, p=0.005), but there were no significant differences in the likelihood of selecting a sugar-sweetened breakfast cereal between participants in the control condition and in the Colombia NNS label condition or between participants in the Chile NNS label condition and in the Colombia NNS label condition (both p>0.05; Figure 4, Figure S2, Table S1).

After seeing all products, among participants who reported noticing any front-of-package labels, those in the Colombia NNS label condition were the most likely to recall seeing labels about NNS (84%, p<0.001 compared to both control and Chile NNS label), followed by those in the Chile NNS label condition (63%, p<0.001 compared to control) and, lastly, by those in the control condition (7%). There were no significant differences between the likelihood of recalling seeing labels about sugar between participants in the control condition and participants in the Colombia NNS label condition (p>0.05), but both were more likely to recall labels about sugar than participants in the Chile NNS label condition (respectively: 95% vs. 89% and 94% vs. 89%, both p<0.001). There were no significant differences between the likelihood of recalling seeing labels about any other nutrients or ingredients between any of the arms (all p>0.05; Figure 5).

**Figure 5.**
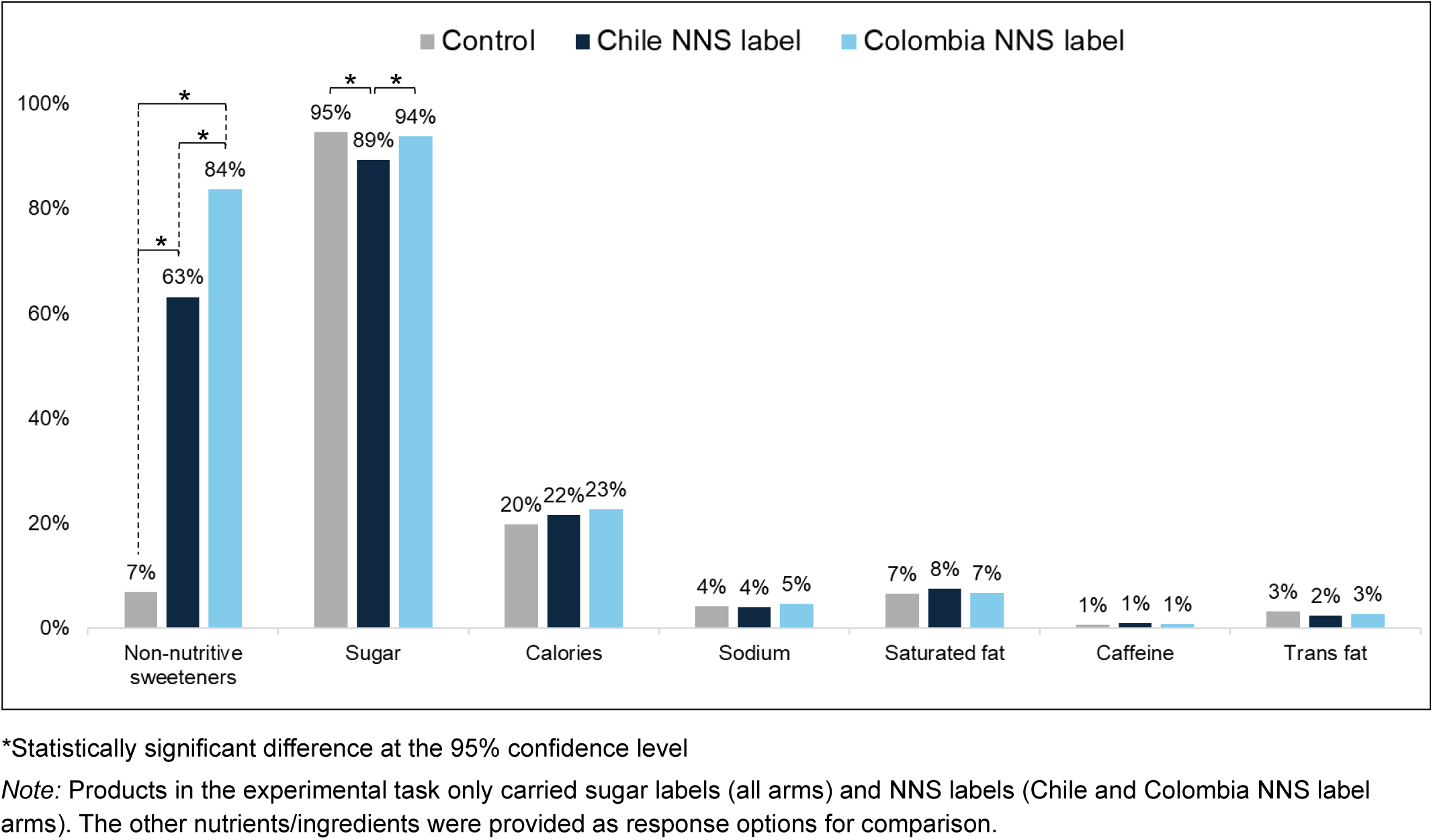
Predicted probability of recalling seeing front-of-package label about each nutrient/ingredient after task 1, by label type (n=3,937)

### Single product assessments

Both types of NNS labels were perceived as more effective than the control label (ADE for Chile NNS label vs. control = 1.53, ADE for Colombia NNS label vs. control = 0.92, both p<0.001), with the Chile NNS label also being perceived as more effective than the Colombia NNS label (ADE=0.61, p<0.001). Both types of NNS labels led to products being perceived as less healthful compared to the control label (ADE for Chile NNS label vs. control = -1.43, ADE for Colombia NNS label vs. control = -0.81, both p<0.001), with the Chile NNS label also leading to products being perceived as less healthful than the Colombia NNS label (ADE = -0.62, p<0.001, Figure 6, Table S2).

**Figure 6.**
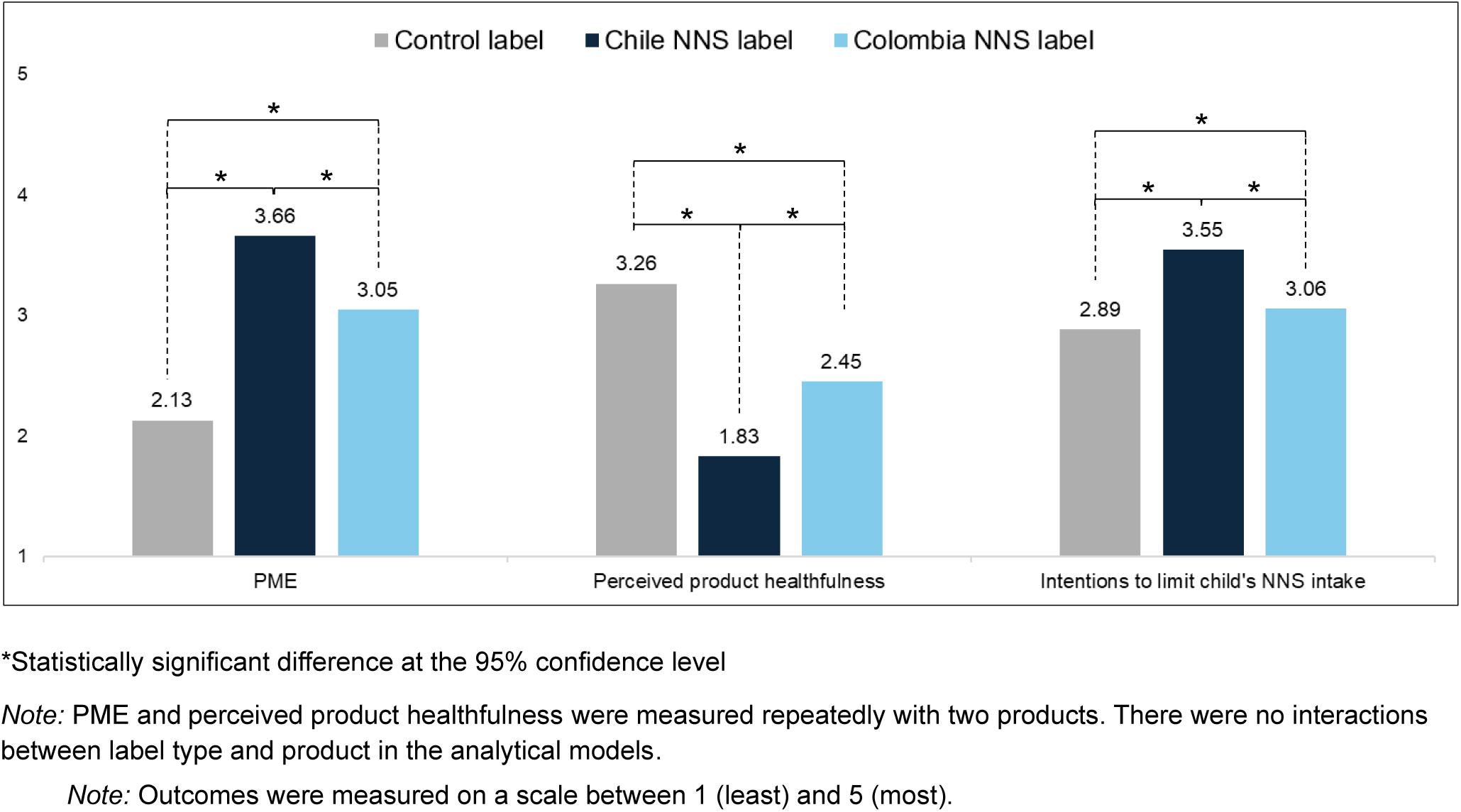
Predicted mean perceived message effectiveness (n=3,503), perceived product healthfulness (n=3,516), and intentions to limit child’s non-nutritive sweetener intake (n=3,514) by label type

Participants in the Chile NNS label condition perceived the NNS-sweetened products seen in this task as the least healthy relative to a version of the same product sweetened with sugar (mean = 2.53; ADE compared to control = -0.71, ADE compared to Colombia NNS label = -0.44, both p<0.001), followed by participants in the Colombia NNS condition (mean = 2.79; ADE compared to control = -0.26, p<0.001), and by participants in the control condition (mean = 3.06; Figure S3, Table S2).

### Intentions to limit NNS

Both types of NNS labels led to higher intentions to limit participants’ children’s NNS intake compared to the control label (ADE for Chile NNS label vs. control = 0.66, ADE for Colombia NNS label vs. control = 0.17, both p<0.001). The Chile NNS label also led to larger intentions to limit one’s child NNS intake than the Colombia NNS label (ADE = 0.49, p<0.001; Figure 6, Table S2).

### Moderation

We found no evidence that gender or educational attainment moderated the effects of the Chile NNS label or the Colombia NNS labels (compared to control) on identification of NNS-containing products, selection of an NNS-sweetened product, or selection of a product without added sweeteners (p>0.05 for all interaction terms). We also found no evidence that NNS tabletop use moderated the effects of the labels on identification of NNS-containing products. Alternatively, we found evidence that NNS tabletop use moderated the effects of the Colombian label (but not of the Chilean label) on selection of an NNS-sweetened product and selection of a product without added sweeteners for one of the three product categories examined (i.e., breakfast cereals, but not fruit drinks or yogurts) – i.e., the effects of the Colombian label on selection of an NNS-sweetened breakfast cereal and selection of a breakfast cereal without added sweeteners were *smaller* among those who reported using tabletop NNS in the last 7 days (ADE=-0.20 and ADE=-0.30, respectively) compared to those who did not report using it (ADE=-0.23 and ADE=-0.37, respectively; p=0.01 for both interaction terms; Tables S4 and S5).

## Discussion

In this online randomized controlled trial with Chilean parents, Chile’s proposed NNS warning label and Colombia’s NNS warning label both greatly increased participants’ ability to identify products containing NNS compared to a control, with the Colombia label exhibiting a slightly larger effect than the Chile label. Both labels also led to reductions in participants’ selection of NNS-sweetened products for their children, with the Chile label exhibiting a slightly larger effect than the Colombia label for one product category. At the same time, both labels led to increases in participants’ selection of products without any added sweeteners for their children, with no differences between the labels. Neither label led to unintended increases in participants’ selection of sugar-sweetened products for their children.

In 2024, a panel of experts recommended NNS warning labels as a potential strategy to address Chilean children’s high NNS intake.^25^ Our findings confirm that such labels can help parents not only identify products containing NNS more easily, but also reduce selection of NNS-sweetened products for their children without increasing selection of sugar-sweetened products. In our study, NNS labels instead led more parents to choose products without added sweeteners, thus achieving their intended goal. We acknowledge that our selection task consistently offered parents options without added sweeteners that were close substitutes for the sugar and NNS-sweetened products offered, and such close substitutes may not always be available in real-world conditions. Nevertheless, our findings suggest that NNS labels hold promise when parents are presented with reasonable alternatives that contain neither sugar nor NNS.

When comparing between the two types of NNS label examined, we found that the Colombian label had a slightly larger effect on participants’ ability to identify products containing NNS and was more readily recalled after the selection task than the Chilean label. These results are likely explained by the Colombian label’s design, which is simpler, more eye-catching, and more familiar to Chileans, thus possibly drawing attention more easily and requiring less cognitive processing. In contrast, when participants were asked to focus on the label, the Chilean label was perceived as more effective at discouraging NNS consumption, led to a greater reduction in the perceived healthfulness of the products to which they were applied, and increased participants’ intentions to limit their children’s NNS intake the most. Yet, during the selection task, both labels performed very similarly. Taken together, these findings suggest that the Chilean label may conceptually hold greater potential for influencing parents’ behavior than the Colombian label, as previously suggested by qualitative studies in Brazil and the US that found the label’s specification that the product was not recommended for children to be particularly compelling to parents.^26,27^ However, the Chilean label’s current visual design may limit its effectiveness in more realistic decision-making contexts. Increasing the size of the Chilean label and/or adding interpretive, attention-grabbing visual elements such as icons may help this label reach its full potential.

Along with gender, we did not find parents’ educational attainment to have an influence on the effects of the NNS labels tested in this study. This is a noteworthy finding considering that children whose mothers have higher educational attainment tend to consume more NNS than children of mothers with lower educational attainment in Chile.^28^ This is a noteworthy finding considering that both the Chilean and the Colombian labels use the term “*edulcorantes*” to refer to NNS, which is a technical term that may be more familiar to individuals with higher literacy levels, and which was only considered the best term to describe NNS by 13% of participants in this study. The two terms that this study’s participants most commonly preferred over “*edulcorantes*” are unfortunately not legally viable for foods labels: “sweeteners” is too vague (also encompassing sugar sweeteners), and “artificial sweeteners” only applies to a subset of NNS (excluding NNS from natural sources such as stevia). Thus, no alternative term appears to be both more comprehensible and as legally viable as “*edulcorantes*.” Yet, the use of this term can pose concerns: in a previous qualitative study in Brazil, participants revealed confusion about the difference between “*edulcorante*” (which is the same in Portuguese and Spanish) and “*corante*” (Portuguese for food colors/dyes).^26^ Given that the Spanish term for colors/dyes, “*colorante*,” is similar, before starting all experimental tasks, we informed participants that the term “*edulcorante*”, which they might encounter during the study, “refers to sweeteners that are not a type of sugar and that have few or no calories.” This explanation may have contributed to the lack of differences in the effects of the NNS labels by participants’ educational attainment, suggesting that, despite the need to use a technical term, NNS labels can still be well understood and influence parents across educational levels similarly if their implementation is accompanied by appropriate education about the label’s content.

This study strengths include its experimental design allowing for causal inference, selection tasks providing participants with several product options developed by a professional designer to mitigate the influence of existing product and brand preferences, a large representation of parents with low educational attainment, and the use of more than one type NNS label for comparison purposes. However, this study also has limitations. As an online study with fictitious products, we cannot establish how generalizable results would be to real-world shopping contexts, where product preferences are already established and substitution options may be more limited. We also used only a few product categories, and thus cannot generalize results to other types of products – however, it is worth noting that we used product categories that have undergone considerable reformulation efforts in Chile and pose some of the main sources of NN intake among children.^11^ Lastly, we used a convenience sample, which limits the generalizability of our findings – yet, previous analyses suggest that online convenience and representative samples tend to produce experimental results similar in direction.^29,30^

## Conclusion

NNS warning labels greatly improved parents’ ability to identify NNS-containing products and led to healthier product choices for their children, showing promise for helping to curb NNS intake in this vulnerable population. The Chilean government should consider increasing the size of its proposed NNS label and/or adding interpretive, attention-grabbing visual elements such as icons to maximize the label’s effectiveness.

## Data Availability

All data produced in the present study will be available in an online repository at the time of publication.

